# Paediatric meningoencephalitis in the molecular diagnostic era: Epidemiological insights from 1,198 suspected cases in Germany between 2016 and 2024

**DOI:** 10.64898/2026.02.15.26346341

**Authors:** Yannik Vollmuth, Benjamin Soric, Jana Beer, Uta Behrends, Marco Paolini, Astrid Blaschek, Melanie Meyer-Bühn, Christoph Klein, Johannes Hübner, Gerhard Dobler, Tilmann Schober

## Abstract

**Background:** The epidemiology of suspected pediatric meningoencephalitis has shifted in the era of conjugate vaccines and multiplex PCR diagnostics, with viral pathogens now predominating over bacterial causes. Updated epidemiologic data are essential to adapt diagnostic and therapeutic algorithms to current clinical practice.

**Methods:** This retrospective single-center study included children and adolescents <18 years who underwent lumbar puncture with cerebrospinal fluid multiplex PCR for suspected central nervous system infection at a tertiary-care pediatric hospital in Germany between 2016 and 2024. Clinical, laboratory, and outcome data were extracted from electronic medical records. Cerebrospinal fluid was analyzed using the BioFire® FilmArray® Meningitis/Encephalitis Panel. Statistical analyses included descriptive statistics, nonparametric group comparisons, receiver operating characteristic analyses.

**Results:** Among 1,198 included children, definite bacterial meningitis was diagnosed in 13 (1.1%), definite viral meningitis in 80 (6.7%), aseptic meningitis of unknown etiology in 131 (11.0%), confirmed/probable encephalitis in 53 (4.4%), and possible encephalitis in 34 (2.8%). Bacterial meningitis accounted for 5.8% of all meningitis cases. A causative pathogen was identified in all bacterial meningitis cases, most commonly *Streptococcus pneumoniae* (n = 7). Enterovirus (n = 52) and parechovirus (n = 9) predominated in viral meningitis, whereas an infectious etiology was identified in only 13 of 53 confirmed/probable encephalitis cases. The Bacterial Meningitis Score showed a sensitivity of 80.0% and a specificity of 57.6%. The recently published UK-ChiMES-pre- and post-lumbar puncture scores demonstrated sensitivities of 84.6% and 76.9% and specificities of 86.3% and 92.7%, respectively.

**Discussion:** Bacterial meningitis was rare in this contemporary cohort, while viral and etiologically unresolved infections predominated despite routine multiplex PCR diagnostics. Clinical prediction scores supported risk stratification, with the UK-ChiMES-pre–lumbar puncture score showing the most favorable balance between sensitivity and specificity and potential to guide diagnostic decisions and antiinfective therapy.

## Background

Acute infections of the central nervous system (CNS), particularly bacterial meningitis, remain medical emergencies associated with substantial morbidity and mortality despite major advances in prevention and treatment [1]. Given the rapid progression and severe consequences of untreated bacterial and selected viral CNS infections, clinicians maintain a low threshold for lumbar puncture in children presenting with symptoms suggestive of meningitis or encephalitis. As a result, many pediatric patients undergo cerebrospinal fluid (CSF) analysis as part of routine diagnostic evaluation.

Over the past decades, the epidemiology of pediatric CNS infections has changed markedly. Epidemiologic data now demonstrate that CNS infections are up to three times more likely to viral than bacterial in origin [2-4]. This shift in the etiologic spectrum is largely attributed to the widespread use of conjugate vaccines, targeting *Neisseria meningitidis, Streptococcus pneumoniae*, and *Haemophilus influenzae* type B, which have substantially reduced the incidence of invasive bacterial disease. Similarly, vaccination against varicella-zoster virus (VZV) has markedly decreased the incidence of VZV-associated CNS infections [2, 5]. Consequently, viral pathogens—most notably herpes simplex virus type 1 (HSV-1), enteroviruses (EV), and, in young infants, human parechoviruses — now predominate [5]. Beyond these established trends, ongoing climatic and ecological changes may further alter the epidemiology of CNS infections, potentially increasing the relevance of emerging or previously rare pathogens even in temperate regions [6].

In parallel, advances in molecular diagnostics have transformed the detection of CNS infections. Multiplex polymerase chain reaction (PCR) assays enable rapid identification of a broad range of pathogens, including EV and echoviruses previously underdiagnosed, thereby increasing diagnostic yield and reshaping the apparent etiologic spectrum of suspected meningoencephalitis [7, 8].

In this study, we aimed to evaluate the contemporary epidemiology of suspected pediatric meningoencephalitis and assess diagnostic strategies including the Bacterial Meningitis Score (BMS) and the recently published UK-ChiMES-pre- and post-lumbar puncture scores in a large, unbiased cohort [5, 9]. We conducted a single-center retrospective study spanning eight years, including all children and adolescents undergoing lumbar puncture for suspected meningitis or encephalitis, to reflect real-world clinical practice.

## Methods

### Study design and patient population

This retrospective, single-center study was conducted at Dr. von Hauner Children’s Hospital, the tertiary-care pediatric hospital of Ludwig Maximilian University (LMU) Munich, Germany. Children and adolescents <18 years of age were eligible if they underwent LP for suspected CNS infection between January 2016 and December 2024 and had cerebrospinal fluid (CSF) multiplex PCR testing performed. Throughout the study period, CSF multiplex PCR was part of routine clinical care for children undergoing LP for suspected CNS infection. Testing was performed using the BioFire® FilmArray® Meningitis/Encephalitis (ME) Panel (BioMérieux, Marcy-l’Étoile, France).

Patients were excluded if they had a ventricular shunt, had undergone recent neurosurgery, or if LP was unsuccessful with no CSF available for analysis. For patients with multiple hospital admissions, only the first admission with LP was included.

### Variables of interest and study definitions

Data were retrospectively extracted from electronic medical records. Demographic, clinical, laboratory, and additional variables are summarized in Tables 1–3 and described in detail in the online Supplementary Methods.

**Table 1.**
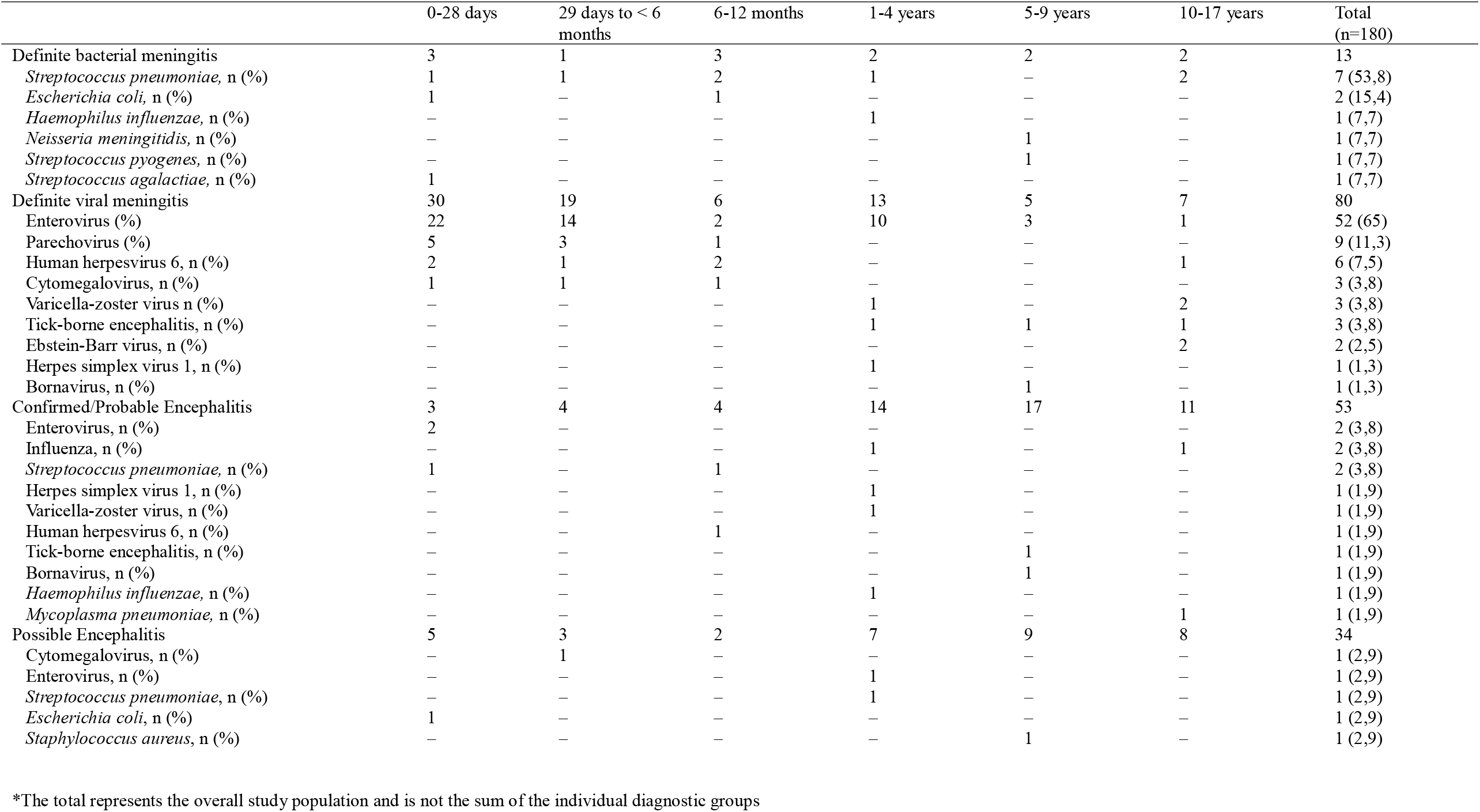
Distribution of specific etiologies of meningitis and encephalitis by age group.

|  | 0-28 days | 29 days to < 6 months | 6-12 months | 1-4 years | 5-9 years | 10-17 years | Total (n=180) |
| --- | --- | --- | --- | --- | --- | --- | --- |
| Definite bacterial meningitis | 3 | 1 | 3 | 2 | 2 | 2 | 13 |
| *Streptococcus pneumoniae*, n (%) | 1 | 1 | 2 | 1 | – | 2 | 7 (53,8) |
| *Escherichia coli*, n (%) | 1 | – | 1 | – | – | – | 2 (15,4) |
| *Haemophilus influenzae*, n (%) | – | – | – | 1 | – | – | 1 (7,7) |
| *Neisseria meningitidis*, n (%) | – | – | – | – | 1 | – | 1 (7,7) |
| *Streptococcus pyogenes*, n (%) | – | – | – | – | 1 | – | 1 (7,7) |
| *Streptococcus agalactiae*, n (%) | 1 | – | – | – | – | – | 1 (7,7) |
| Definite viral meningitis | 30 | 19 | 6 | 13 | 5 | 7 | 80 |
| Enterovirus (%) | 22 | 14 | 2 | 10 | 3 | 1 | 52 (65) |
| Parechovirus (%) | 5 | 3 | 1 | – | – | – | 9 (11,3) |
| Human herpesvirus 6, n (%) | 2 | 1 | 2 | – | – | 1 | 6 (7,5) |
| Cytomegalovirus, n (%) | 1 | 1 | 1 | – | – | – | 3 (3,8) |
| Varicella-zoster virus n (%) | – | – | – | 1 | – | 2 | 3 (3,8) |
| Tick-borne encephalitis, n (%) | – | – | – | 1 | 1 | 1 | 3 (3,8) |
| Ebstein-Barr virus, n (%) | – | – | – | – | – | 2 | 2 (2,5) |
| Herpes simplex virus 1, n (%) | – | – | – | 1 | – | – | 1 (1,3) |
| Bornavirus, n (%) | – | – | – | – | 1 | – | 1 (1,3) |
| Confirmed/Probable Encephalitis | 3 | 4 | 4 | 14 | 17 | 11 | 53 |
| Enterovirus, n (%) | 2 | – | – | – | – | – | 2 (3,8) |
| Influenza, n (%) | – | – | – | 1 | – | 1 | 2 (3,8) |
| *Streptococcus pneumoniae*, n (%) | 1 | – | 1 | – | – | – | 2 (3,8) |
| Herpes simplex virus 1, n (%) | – | – | – | 1 | – | – | 1 (1,9) |
| Varicella-zoster virus, n (%) | – | – | – | 1 | – | – | 1 (1,9) |
| Human herpesvirus 6, n (%) | – | – | 1 | – | – | – | 1 (1,9) |
| Tick-borne encephalitis, n (%) | – | – | – | – | 1 | – | 1 (1,9) |
| Bornavirus, n (%) | – | – | – | – | 1 | – | 1 (1,9) |
| *Haemophilus influenzae*, n (%) | – | – | – | 1 | – | – | 1 (1,9) |
| *Mycoplasma pneumoniae*, n (%) | – | – | – | – | – | 1 | 1 (1,9) |
| Possible Encephalitis | 5 | 3 | 2 | 7 | 9 | 8 | 34 |
| Cytomegalovirus, n (%) | – | 1 | – | – | – | – | 1 (2,9) |
| Enterovirus, n (%) | – | – | – | 1 | – | – | 1 (2,9) |
| *Streptococcus pneumoniae*, n (%) | – | – | – | 1 | – | – | 1 (2,9) |
| *Escherichia coli*, n (%) | 1 | – | – | – | – | – | 1 (2,9) |
| *Staphylococcus aureus*, n (%) | – | – | – | – | 1 | – | 1 (2,9) |

\*The total represents the overall study population and is not the sum of the individual diagnostic groups

**Table 2.** Baseline characteristics according to diagnostic classification.

|  | Definite bacterial meningitis | Definite viral meningitis | Aseptic meningitis with unknown etiology | Confirmed/probable encephalitis | Possible encephalitis | Diagnosis of febrile convulsions | CNS Lyme disease | Control group | Total* |
| --- | --- | --- | --- | --- | --- | --- | --- | --- | --- |
| n | 13 | 80 | 131 | 53 | 34 | 132 | 23 | 902 | 1198 |
| Sex, n (%) |  |  |  |  |  |  |  |  |  |
| Female | 5 (38.5) | 34 (42.5) | 57 (43.5) | 23 (43.4) | 16 (47.1) | 63 (47.7) | 7 (30.4) | 407 (45.1) | 548 (45.7) |
| Male | 8 (61.5) | 46 (57.5) | 74 (56.5) | 30 (56.6) | 18 (52.9) | 69 (52.3) | 16 (69.6) | 495 (54.9) | 650 (54.3) |
| Age Groups, n (%) |  |  |  |  |  |  |  |  |  |
| 0-28 days | 3 (23.1) | 30 (37.5) | 22 (16.8) | 3 (5.7) | 5 (14.7) | 1 (0.8) | 0 (0.0) | 186 (20.6) | 246 (20.5) |
| 29 days to < 6 months | 1 (7.7) | 19 (23.8) | 18 (13.7) | 4 (7.5) | 3 (8.8) | 1 (0.8) | 0 (0.0) | 101 (11.2) | 140 (11.7) |
| 6-12 months | 3 (23.1) | 6 (7.5) | 7 (5.3) | 4 (7.5) | 2 (5.9) | 19 (14.4) | 0 (0.0) | 47 (5.2) | 67 (5.6) |
| 1-4 years | 2 (15.4) | 13 (16.3) | 15 (11.5) | 14 (26.4) | 7 (20.6) | 85 (64.4) | 1 (4.3) | 186 (20.6) | 241 (20.1) |
| 5-9 years | 2 (15.4) | 5 (6.3) | 28 (21.4) | 17 (32.1) | 9 (26.5) | 23 (17.4) | 11 (47.8) | 123 (13.6) | 175 (14.6) |
| 10-17 years | 2 (15.4) | 7 (8.8) | 41 (31.3) | 11 (20.8) | 8 (23.5) | 3 (2.3) | 11 (47.8) | 259 (28.7) | 329 (27.5) |
| Past medical history, n (%) |  |  |  |  |  |  |  |  |  |
| Immunocompromised | 1 (7.7) | 3 (3.8) | 20 (15.3) | 6 (11.3) | 2 (5.9) | 0 (0.0) | 0 (0.0) | 47 (5.2) | 73 (6.1) |
| Malignancy | 0 (0.0) | 0 (0.0) | 11 (8.4) | 1 (1.9) | 1 (2.9) | 0 (0.0) | 0 (0.0) | 34 (3.8) | 45 (3.8) |
| Previous neurological condition | 0 (0.0) | 2 (2.5) | 11 (8.4) | 15 (28.3) | 8 (23.5) | 7 (5.3) | 2 (0.0) | 74 (8.2) | 110 (9.2) |
| Vaccines up to date, n (%) |  |  |  |  |  |  |  |  |  |
| Yes | 10 (76.9) | 69 (86.3) | 91 (69.5) | 40 (75.5) | 24 (70.6) | 97 (73.5) | 18 (78.3) | 628 (69.6) | 853 (71.2) |
| No | 3 (16.7) | 3 (3.8) | 6 (4.6) | 4 (7.5) | 3 (8.8) | 11 (8.3) | 3 (13.0) | 54 (6.0) | 77 (6.4) |
| Not reported | 0 (0.0) | 8 (10.0) | 34 (26.0) | 9 (17.0) | 7 (20.6) | 24 (18.2) | 2 (8.7) | 220 (24.4) | 268 (22.4) |
| History, n (%) |  |  |  |  |  |  |  |  |  |
| Symptom duration (days) before hospital presentation (median, IQR) | 1.0 (0.0-3.0) | 0.0 (0.0-1.0) | 1.0 (0.0-6.0) | 1.0 (0.0-3.0) | 1.0 (0.0-3.0) | 0.0 (0.0-1.0) | 3.0 (0.0-17.0) | 1.0 (0.0-4.0) | 1 (0.0-3.0) |

\*The total represents the overall study population and is not the sum of the individual diagnostic groups

**Table 3.** Clinical characteristics and outcomes according to diagnostic classification.

|  | Definite bacterial meningitis | Definite viral meningitis | Aseptic meningitis with unknown etiology | Confirmed/ Probable encephalitis | Possible encephalitis | Diagnosis of febrile convulsions | CNS Lyme disease | Control group | Total* |
| --- | --- | --- | --- | --- | --- | --- | --- | --- | --- |
| n | 13 | 80 | 131 | 53 | 34 | 132 | 23 | 902 | 1198 |
| Hospital-acquired, n (%) | 0 (0.0) | 0 (0.0) | 0 (0.0) | 1 (1.9) | 2 (5.9) | 0 (0.0) | 0 (0.0) | 4 (0.4) | 7 (0.6) |
| Community-acquired, n (%) | 13 (100.0) | 80 (0.0) | 131 (0.0) | 52 (0.98) | 32 (94.1) | 132 (100.0) | 23 (100.0) | 898 (99.6) | 1191 (99.4) |
| Symptoms, n (%) |  |  |  |  |  |  |  |  |  |
| Fever | 10 (76.9) | 65 (81.3) | 48 (36.6) | 37 (69.8) | 20 (58.8) | 132 (100.0) | 3 (13.0) | 310 (34.4) | 495 (41.3) |
| Headache | 3 (23.1) | 14 (17.5) | 34 (26.0) | 13 (24.5) | 5 (14.7) | 5 (3.8) | 12 (52.2) | 164 (18.2) | 225 (18.8) |
| Neck stiffness | 6 (46.2) | 10 (12.5) | 10 (7.6) | 6 (11.3) | 4 (11.8) | 8 (6.1) | 2 (8.7) | 39 (4.3) | 78 (6.5) |
| Irritability | 4 (30.8) | 23 (28.7) | 19 (14.5) | 13 (24.5) | 13 (38.2) | 34 (25.8) | 0 (0.0) | 128 (14.2) | 193 (16.1) |
| Rash | 4 (30.8) | 20 (25.0) | 10 (7.6) | 1 (1.9) | 2 (5.9) | 15 (11.4) | 3 (13.0) | 54 (6.0) | 99 (8.3) |
| Vomiting | 7 (53.8) | 21 (26.3) | 27 (20.6) | 24 (45.3) | 11 (32.4) | 33 (25.0) | 3 (13.0) | 150 (16.6) | 230 (19.2) |
| Bulging fontanelle | 0 (0.0) | 1 (1.3) | 1 (0.8) | 1 (1.9) | 0 (0.0) | 1 (0.8) | 0 (0.0) | 3 (0.3) | 10 (0.8) |
| Altered level of consciousness >24h | 4 (30.8) | 11 (13.8) | 23 (17.6) | 53 (100.0) | 34 (100.0) | 12 (9.1) | 0 (0.0) | 39 (4.3) | 136 (11.4) |
| Personality change >24h | 0 (0.0) | 2 (2.5) | 17 (13.0) | 13 (24.5) | 3 (8.8) | 2 (1.5) | 2 (8.7) | 52 (5.8) | 73 (6.1) |
| Seizure <72h before or during hospital stay | 3 (23.1) | 11 (13.8) | 27 (27.4) | 38 (71.7) | 19 (55.9) | 132 (100.0) | 0 (0.0) | 233 (25.8) | 328 (27.4) |
| Outcome, n (%) |  |  |  |  |  |  |  |  |  |
| Length of stay (median, IQR) | 14.0 (11.5-26.0) | 5.0 (4.0-7.0) | 7.0 (5.0-13.0) | 11.0 (5.0-24.0) | 8.0 (4.0-17.3) | 3.0 (2.3-4.0) | 5.0 (3.0-11.0) | 5.0 (3.0-8.0) | 4.0 (3.0-7.0) |
| ICU stay | 7 (53.8) | 13 (16.3) | 44 (33.6) | 46 (86.8) | 24 (70.6) | 29 (22.0) | 1 (4.3) | 230 (25.5) | 343 (28.6) |
| Death | 0 (0.0) | 1 (1.3) | 0 (0.0) | 2 (3.8) | 0 (0.0) | 0 (0.0) | 0 (0.0) | 5 (0.6) | 9 (0.8) |

\*The total represents the overall study population and is not the sum of the individual diagnostic groups

Meningitis was classified according to predefined criteria adapted from the UK-ChiMES network, based on clinical features, CSF findings and microbiological results [5]. CSF pleocytosis was defined as a CSF white blood cell (WBC) count ≥20 × 10□/L in neonates aged 0–28 days and >5 × 10□/L in children > 28 days, with correction for blood contamination using a ratio of 500 × 10□/L red blood cells (RBC) to 1 × 10□/L WBC [5]. Definite bacterial meningitis was defined by identification of a relevant bacterial pathogen in CSF, or by CSF pleocytosis with detection of a relevant bacterial pathogen in blood by culture or PCR, or by a positive CSF gram staining with a corresponding blood pathogen. Relevant pathogens included *Neisseria meningitidis, Streptococcus pneumoniae, Haemophilus influenzae*, Group B *Streptococcus, Escherichia coli, Staphylococcus aureus*, and Group A *Streptococcus*. Definite viral meningitis was defined by CSF pleocytosis with detection of a viral pathogen in CSF or blood, or by detection of enterovirus, parechovirus, or herpes simplex virus (HSV) in CSF irrespective of CSF WBC count. Aseptic meningitis with unknown etiology was defined as CSF pleocytosis without pathogen identification [5]. (Supplementary Figure (Fig) 1 and Supplementary Table 2)

Encephalitis was classified according to the International Encephalitis Consortium consensus criteria [10]. The required major criterion was altered mental status, lethargy or personality change lasting ≥24 hours without an alternative cause. Minor criteria included fever ≥38 °C within 72 hours of presentation, seizures not attributable to a preexisting disorder, new-onset focal neurological deficits, CSF WBC count ≥5 cells/mm^3^, neuroimaging findings suggestive of encephalitis, or electroencephalographic abnormalities consistent with encephalitis. Cases were categorized as possible or probable encephalitis based on the number of fulfilled minor criteria [10] (Supplementary Fig 1; Supplementary Table 2). Cases of probable encephalitis, with additional microbiological confirmation of a causative pathogen were defined as confirmed encephalitis.

Risk stratification for bacterial meningitis was evaluated within the study cohort using established prediction tools. The BMS was applied to children aged ≥29 days with CSF pleocytosis. Patients were classified as being at very low risk for bacterial meningitis if none of the following criteria were present and as not very low risk if one or more criteria were present: positive CSF gram staining, CSF absolute neutrophil count ≥1000 cells/µL, CSF protein concentration ≥80 mg/dL, peripheral blood absolute neutrophil count ≥10 000 cells/µL, or a history of seizure before or at the time of presentation [9, 11, 12] (Supplementary Table 3). The pre- and post-lumbar puncture (Pre-LP and Post-LP) points-based score described by the UK-ChiMES study group [4] was applied to all eligible patients and assessed in the cohort. This score incorporates clinical features and laboratory parameters obtained before and after LP, including vomiting, altered consciousness, bulging fontanelle, peripheral lymphocyte count, C-reactive protein (CRP) levels, rash, CSF glucose, CSF protein, CSF WBC count, and CSF neutrophil count, using predefined cut-off values (Supplementary Table 4).

### Data analysis

Data collection and the creation of tables and figures were performed using Microsoft Excel, Microsoft Word, and Microsoft PowerPoint (Microsoft Corp., Redmond, WA, USA), as well as BioRender (BioRender, Toronto, ON, Canada). Statistical analyses were performed using SPSS (IBM Corp., Armonk, NY, USA). Descriptive statistics were used to summarize the data. Categorical variables are reported as frequencies and percentages, and continuous variables as medians with interquartile ranges (IQR). Receiver operating characteristic (ROC) curve analysis was performed where applicable. A two-sided p value of <0.05 was considered statistically significant.

The study approved by the Ethics Committee of the LMU Munich (project number 24-1003) and conducted in accordance with the Declaration of Helsinki. The Strengthening the Reporting of Observational studies in Epidemiology (STROBE) guidelines were followed (see corresponding checklist, Supplemental Material) [13].

## Results

A total of 1,348 children undergoing lumbar puncture with multiplex PCR were initially assessed for eligibility (Fig 1). After excluding 71 patients with ventricular shunt or recent neurosurgery, 37 patients without CSF analysis, and 42 repeat admissions, 1,198 children were included in the final analysis.

**Figure 1.**
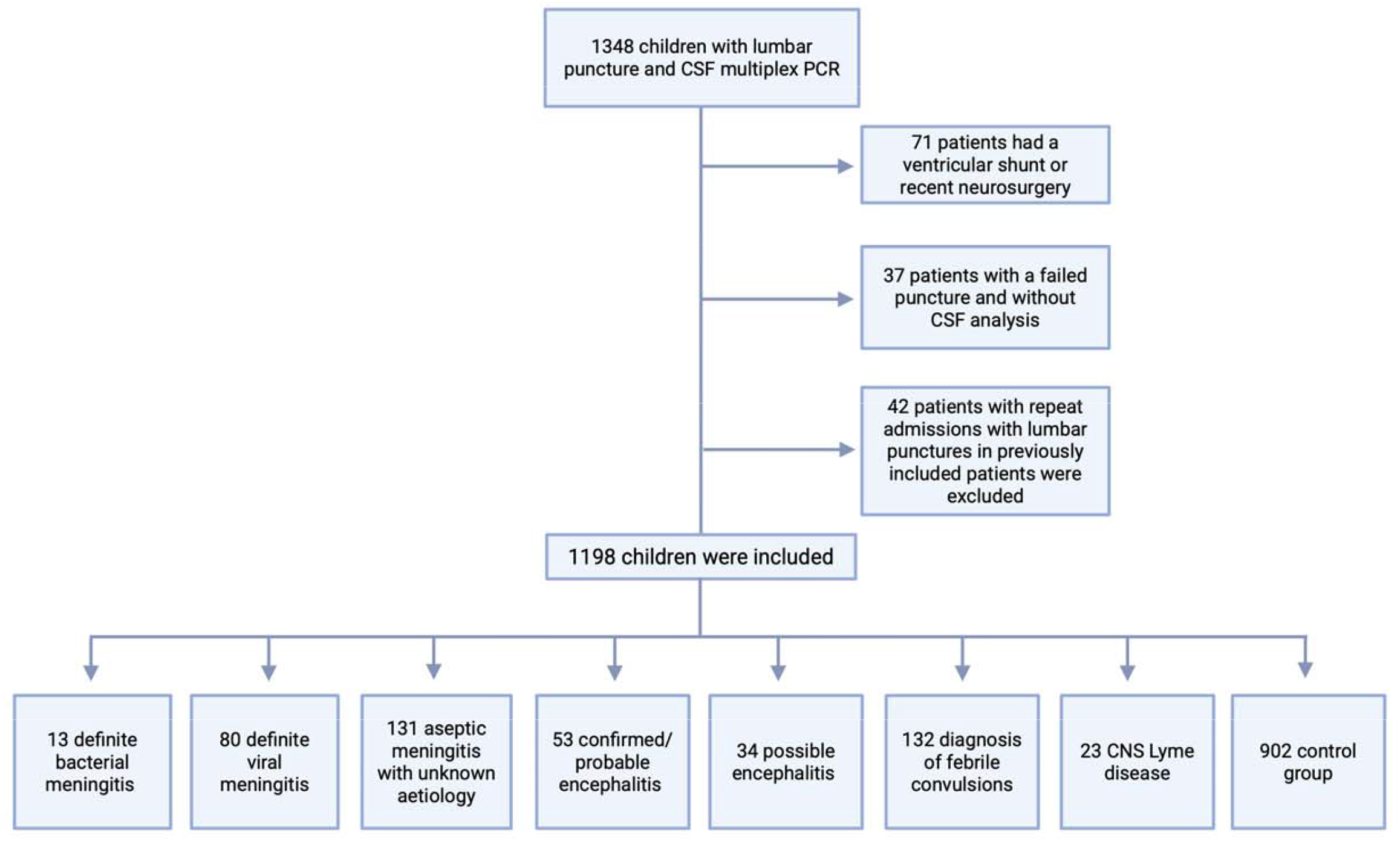
Flowchart of hospitalized children <18 years of age with suspected CNS infection between January 2016 and December 2024

### Clinical epidemiology

According to the predefined diagnostic classification (Supplementary Fig 1; Supplementary Table 2), the cohort comprised 13 (1.1%) patients with definite bacterial meningitis, 80 (6.7%) with definite viral meningitis, 131 (11.0 %) with aseptic meningitis of unknown etiology, 53 (4.4%) with confirmed/probable encephalitis, 34 (2.8%) with possible encephalitis, 132 (11.0%) with a diagnosis of febrile convulsions, 23 (1.9%) with CNS Lyme disease, and 902 (75,3%) children assigned to the control group (Figure 1). When combining definite bacterial, definite viral, and aseptic meningitis of unknown etiology, a total of 224 meningitis cases were identified. Of these, 13 cases were classified as definite bacterial meningitis, corresponding to 5.8% (13/224) of all meningitis cases. Aseptic meningitis of unknown etiology accounted for 58.5% (131/224) of all meningitis diagnoses.

In patients with definite bacterial meningitis (n = 13), a causative pathogen was identified in all cases, with *Streptococcus pneumoniae* representing the most frequent etiology (7/13), followed by Escherichia coli (2/13), while *Haemophilus influenzae, Neisseria meningitidis, Streptococcus pyogenes* and *Streptococcus agalactiae* were each detected in single cases. In definite viral meningitis (n = 80), EV was the predominant pathogen across all age groups, accounting for 52 cases, followed by parechovirus (9 cases) and human herpesvirus 6 (HHV6) (6 cases); other viral pathogens were detected less frequently (Table 1).

In contrast, among patients with probable encephalitis (n = 53), a specific infectious etiology was identified in only 24.5% (13/53) cases; these cases were reclassified as confirmed encephalitis. (Table 1). Detected pathogens included EV and influenza virus (each 2 cases), *Streptococcus pneumoniae* (2 cases), as well as HSV-1, VZV, HHV6, tick-borne encephalitis (TBE) virus, bornavirus, *Haemophilus influenzae* and *Mycoplasma pneumoniae* (each 1 case). Consequently, the majority of confirmed/probable encephalitis cases remained without an identified pathogen despite multiplex PCR testing and additional infectious workup. A similar pattern was observed in possible encephalitis (n = 34), in which 85.3% (29/34) of patients had no detectable infectious etiology (Table 1).

Baseline demographic and clinical characteristics of the study population stratified by diagnostic category are summarized in Table 2. Overall, 20.5% (246/1198) of the patients were neonates and 17.3% (207/1198) were infants beyond neonatal age. Clinical presentation differed substantially across diagnostic categories as shown in Table 3. Fever was present in 76.9% (10/13) of patients with definite bacterial meningitis, 81.3% (65/80) with definite viral meningitis, 69.8% (37/53) with confirmed/probable encephalitis and 58.8% (20/34) with possible encephalitis, compared with 34.4% (310/902) in the control group. Altered level of consciousness lasting longer than 24 hours was reported in all patients with confirmed/probable or possible encephalitis, compared with 30.8% (4/13) in bacterial meningitis and 13.8% (11/80) in viral meningitis. Seizures before or during hospitalization occurred in 71.7% (38/53) of patients with confirmed/probable encephalitis and 55.9% (19/34) with possible encephalitis, whereas they were less frequent in meningitis groups and occurred in 23.1% (3/13) of definite bacterial meningitis and 13.8% (11/80) of viral meningitis. Admission to the intensive care unit (ICU) occurred in 53.8% (7/13) of patients with bacterial meningitis and 86.8% (46/53) with confirmed/probable encephalitis. Overall mortality was low, with deaths reported in 0.8% (9/1198) of the total study population (Table 3), including none of the patients with a bacterial meningitis.

### Evaluation of biomarkers

Patients with definite bacterial meningitis showed substantially higher inflammatory markers in blood than all other groups (Table 4). Median absolute neutrophil counts were highest in definite bacterial meningitis, while lymphocyte counts were lower compared with viral and aseptic meningitis. Blood CRP and procalcitonin (PCT) concentrations were highest in bacterial meningitis, with median CRP levels of 126 mg/L (IQR 64–220) and PCT levels of 7.8 ng/mL (IQR 3.6–21.2), while remaining low in non-bacterial diagnostic categories. Median serum interleukin-6 (IL-6) levels were markedly elevated in definite bacterial meningitis (976 pg/mL, IQR 87.0–3884) and substantially lower in definite viral meningitis (46.8 pg/mL, IQR 17.6–103.0) and aseptic meningitis of unknown etiology (68.3 pg/mL, IQR 13.1–195.0). Receiver operating characteristic analyses assessed the discriminatory performance of inflammatory biomarkers for definite bacterial meningitis (Supplementary Fig 2; Supplementary Table 5). Areas under the curve were 0.919 (95% CI 0.859–0.979) for CRP, 0.852 (95% CI 0.714–0.990) for PCT and, 0.782 (95% CI 0.641–0.923) for IL-6, all statistically significant (p < 0.001). For CRP, an optimal cut-off value of ≥50.7 mg/L yielded a sensitivity of 84.6% and a specificity of 85.9%, %, corresponding to a Youden index of 0.705. PCT demonstrated a sensitivity of 90.0% and a specificity of 85.8% at a cut-off of ≥3.35 ng/mL, with a Youden index of 0.758. IL-6 showed a sensitivity of 60% and a specificity of 90.5% at a cut-off of ≥949 pg/mL, resulting in a Youden index of 0.505.

**Table 4.** Laboratory parameters according to diagnostic classification.

|  | Definite bacterial meningitis | Definite viral meningitis | Aseptic meningitis with unknown aetiology | Confirmed/ Probable encephalitis | Possible encephalitis | Diagnosis of febrile convulsions | CNS Lyme disease | Control group | Total |
| --- | --- | --- | --- | --- | --- | --- | --- | --- | --- |
| n | 13 | 80 | 131 | 53 | 34 | 132 | 23 | 755 | 1198 |
| Blood Parameters (median, IQR) |  |  |  |  |  |  |  |  |  |
| WBC absolute (G/L) § | 17.3 (11.8-23.7) | 8.9 (6.3-11.1) | 9.9 (7.0-15.9) | 11.4 (7.5-15.3) | 10.0 (6.5-13.2) | 9.4 (6.2-14.1) | 8.0 (7.0-9.4) | 9.0 (6.4-14.1) | 9.0 (6.4-13.3) |
| Absolute neutrophil count (G/L)* | 10.4 (6.7-18.0) | 4.0 (2.6-6.0) | 5.1 (3.0-10.4) | 7.4 (4.3-11.3) | 5.8 (2.6-9.7) | 6.7 (3.9-9.5) | 4.5 (3.6-5.8) | 4.6 (2.7-8.4) | 4.8 (2.8-8.5) |
| Absolute lymphocyte count (G/L)* | 2.0 (1.4-7.1) | 2.9 (1.6-4.2) | 2.8 (1.7-4.6) | 2.1 (1.3-2.9) | 1.9 (1.3-3.6) | 1.9 (1.2-2.6) | 2.7 (2.2-3.2) | 2.3 (1.4-3.6) | 2.6 (1.7-4.0) |
| CRP (mg/L)+ | 126.0 (64.0-220.0) | 6 (2.0-19.0) | 8.0 (1.0-50.0) | 10.0 (1.0-36.0) | 7.0 (2.0-47.0) | 9.0 (3.0-21.0) | 1.0 (1.0-1.0) | 3.0 (1.0-32.0) | 4.0 (1.0-27.0) |
| PCT (ng/mL)++ | 7.8 (3.6-21.2) | 0.1 (0.1-0.3) | 0.2 (0.1-1.6) | 0.2 (0.1-1.8) | 0.3 (0.1-3.2) | 0.3 (0.1-0.7) | 0.1 (0.1-0.1) | 0.2 (0.1-1.2) | 0.2 (0.1-1.1) |
| IL-6 (pg/mL)+++ | 976 (87.0-3884) | 46.8 (17.6-103.0) | 68.3 (13.1-195.0) | 32.6 (8.1-178.0) | 51.1 (10.6-191.8) | 57.0 (28.9-137.0) | n/a | 44.6 (13.2-178.0) | 46.2 (13.0-168.0) |
| CSF Parameters (median, IQR) |  |  |  |  |  |  |  |  |  |
| Corrected CSF WBC count | 293 (73.5-2029.5) | 6.0 (1.0-104.0) | 24.0 (11.0-73.0) | 5.0 (1.0-65.0) | 0.9 (0.0-3.0) | 1.0 (0.0-2.0) | 54.0 (15.8-174.9) | 1.0 (0.5-3.0) | 1.0 (0.7-4.0) |
| CSF Segmented neutrophilic granulocytes (%)# | 72.0 (47.0-88.3) | 32.0 (5.0-62.3) | 30.0 (5.0-62.0) | 38.5 (10.8-78.5) | 5.0 (1.5-24.0) | 52.5 (10.25-63.0) | 1.0 (1.0-2.75) | 32.0 (7.5-57.0) | 32.0 (5.0-57.3) |
| CSF Lymphocytes (%)# | 17.0 (6.0-39.0) | 34.0 (14.0-56.0) | 65.0 (20.0-86.5) | 57.0 (19.0-84.0) | 50.0 (23.0-71.5) | 45.5 (34.0-81.0) | 86.5 (83.0-89.0) | 32.0 (15.8-66.0) | 36.0 (17.0-76.0) |
| CSF Monocytes (%)# | 6.5 (3.0-12.3) | 14.0 (5.5-43.5) | 11.0 (5.0-18.5) | 9.0 (3.5-13.0) | 11.5 (4.5-28.5) | 6.0 (2.0-17.0) | 7.0 (4.8-9.0) | 16.0 (6.0-36.0) | 11.0 (5.0-30.0) |
| Corrected CSF Protein (mg/dL) | 96.7 (35.0-220.2) | 43.6 (28.6-62.9) | 45.0 (26.3-91.8) | 35.3 (18.2-88.7) | 27.0 (15.0-45.2) | 15.0 (12.5-19.0) | 50.0 (28.0-73.7) | 24.0 (17.0-48.0) | 26.0 (17.0-52.0) |
| CSF Glucose (mg/dL) | 43.0 (2.0-71.4) | 52.5 (45.6-61.0) | 56.1 (50.4-64.5) | 65.8 (53.3-82.7) | 64.7 (56.3-76.1) | 68.0 (61.6-76.9) | 57.5 (49.3-66.5) | 57.9 (52.3-65.0) | 58.0 (51.7-65.6) |
| CSF Lactate (mmol/L) | 2.9 (2.0-8.3) | 1.6 (1.3-1.9) | 1.7 (1.5-2.2) | 1.8 (1.5-2.5) | 1.6 (1.4-2.2) | 1.5 (1.3-1.7) | 1.7 (1.5-2.1) | 1.5 (1.35-1.7) | 1.51 (1.4-1.8) |

§ For absolute WBC count, data were missing for a total of 42 patients across all categories.
\* For leucocyte subgroups, data were missing for a total of 63 patients across all categories.
+ For CRP, data were missing for a total of 62 patients across all categories.
++ For PCT, data were missing for a total of 710 patients across all categories.
+++ For IL-6, data were missing for a total of 638 patients across all categories.

### Performance of meningitis prediction scores

The diagnostic performance of the BMS as well as the recently published pre–lumbar puncture (UK-ChiMES-pre-LP) and post–lumbar puncture (UK-ChiMES-post-LP) scores was evaluated in the overall study population (Table 5) [5, 14, 15]. The BMS showed a sensitivity of 80.0% and a specificity of 57.6%, with a negative predictive value (NPV) of 99.6% and a positive predictive value (PPV) of 2.0%. The UK-ChiMES-pre-LP score demonstrated a sensitivity of 84.6% and a specificity of 86.3%, with a NPV of 99.8% and a PPV of 6.4%. The UK-ChiMES-post-LP score yielded a sensitivity of 76.9% and a specificity of 92.7%, with a NPV of 99.7% and a PPV of 10.3% (Table 5). Among the 13 probable cases of bacterial meningitis, several were not identified by one or more risk scores. Individual score charts and patient characteristics are shown in Supplementary Tables 6 and 7. Among those, 3 cases were not considered to be a meningitis by the treating physicians, including one case each missed by the BMS score and the UK-ChiMES-pre-LP and post-LP score.

**Table 5.** Evaluation of the Bacterial Meningitis Score and UK-ChiMES Pre/Post-LP Score.

| Measure | Bacterial meningitis Score | UK-ChiMES Pre-LP Score | UK-ChiMES Post-LP Score |
| --- | --- | --- | --- |
| Sensitivity | 80.0% | 84.6% | 76.9% |
| Specificity | 57.6% | 86.3% | 92.7% |
| Positive Predictive Value (PPV) | 2.0% | 6.4% | 10.3% |
| Negative Predictive Value (NPV) | 99.6% | 99.8% | 99.7% |
| Accuracy | 57.9% | 86.3% | 86.3% |
| Positive Likelihood Ratio | 1.89 | 6.17 | 10.5 |
| Negative Likelihood Ratio | 0.35 | 0.18 | 0.25 |

## Discussion

In this retrospective single-center cohort, we investigated the contemporary clinical and epidemiologic spectrum of suspected meningoencephalitis in children in the era of conjugate vaccination and routine multiplex PCR diagnostics. We demonstrate a predominance of viral pathogens and a substantial proportion of cases remaining etiologically unresolved despite advanced diagnostics. Clinical prediction scores showed good NPV, supporting their use as rule-out tools, but low PPV reflecting the very low prevalence of bacterial meningitis in this cohort.

The most important finding of this study is the low proportion of bacterial meningitis observed in a large unselected cohort of children undergoing LP for suspected CNS infection. Among 1,198 patients, only 13 (1.1%) cases of definite bacterial meningitis were identified, corresponding to a number needed to test (NNT) of 92 LP. When restricting the analysis to children with meningitis, bacterial etiologies accounted for 5.8% of cases, placing our findings at the lower end of proportions reported in previous studies from Europe and North America (4–18%). [5, 9, 14, 15]. Aseptic meningitis of unknown etiology accounted for 58.5% of all meningitis cases, consistent with prior reports in which no causative pathogen was identified in 30–76% of children. [5, 16, 17]. Variability across studies likely reflects differences in population selection, diagnostic strategies, and study design. Our pragmatic inclusion of all hospitalized children evaluated for suspected meningitis or encephalitis therefore provides a representative reflection of real-world clinical practice. *Streptococcus pneumoniae* was the most frequently identified bacterial pathogen, consistent with earlier reports from the United States. In contrast, recent data from the United Kingdom identified *Neisseria meningitidis* as the predominant cause, reflecting a higher meningococcal incidence that is also observed in public health surveillance data. [5, 18-20]

In contrast, viral meningitis was substantially more frequent, especially EV and human parechoviruses. This is consistent with the published literature, including large national surveillance studies from the United Kingdom and Ireland, which report that EV account for approximately 75– 95% of laboratory-confirmed viral meningitis cases in young infants, while human parechoviruses contribute 5–12%, particularly among infants younger than 3–6 months of age. These proportions correspond to incidences of 0.79 and 0.04 per 1,000 live births, respectively. [21, 22] Similarly, international data estimate an annual burden of approximately 75,000 cases of enteroviral meningitis in the United States, predominantly affecting neonates and young infants [23-25].

Despite routine use of multiplex RPC and additional diagnostics, the majority of encephalitis cases still lack an identified infectious source. This observation is consistent with a comprehensive review reporting that over 50% of pediatric encephalitis cases remained of unknown etiology [26]. Similarly, the International Encephalitis Consortium concluded that, in routine clinical practice, a specific etiology is identified in fewer than 50% of encephalitis cases, despite advanced diagnostics [10]. These findings emphasize the ongoing diagnostic gap and the need for further approaches, such as sequencing, to improve identification.

Rising rates of TBE have been described and linked to environmental and ecological factors, including climate-driven increases in tick activity and shifts in animal host distribution that enhance virus circulation and human exposure [27]. In Germany, TBE incidence is approximately 1–2 cases per 100,000 inhabitants, with higher and increasing rates in endemic regions in southern Germany, including the hospital catchment area [28]. In contrast, the incidence of Lyme borreliosis ranges from approximately 40 to over 200 cases per 100,000 inhabitants. Neurological manifestations occur in only 3–15% of infections [29]. In our cohort, three cases of tick-borne encephalitis (TBE) were identified, possibly reflecting underdiagnosis due to its often nonspecific clinical presentation, as testing was performed only when there was specific clinical suspicion and not routinely in all cases. In contrast, 23 cases of neuroborreliosis were detected, likely facilitated by characteristic features such as facial nerve palsy, which increase diagnostic suspicion.

In our cohort, the BMS showed a high NPV, confirming its usefulness as a rule-out tool, but limited specificity and low PPV of only 2% substantially restricted its ability to identify children at high risk of bacterial meningitis. In contrast, both the UK-ChiMES-pre-LP and post-LP scores demonstrated improved overall diagnostic performance, with higher specificity while maintaining excellent NPV. This manuscript represents the first independent evaluation of these scores. Their performance was broadly comparable to the original description by Martin et al., who reported sensitivities of approximately 82% and 84% and specificities of 71% and 93% for the pre-LP and post-LP models, respectively [5]. Despite being applied before lumbar puncture, the UK-ChiMES-pre-LP score outperformed the Bacterial Meningitis Score, achieving higher sensitivity and substantially improved specificity, with fewer false-positive classifications. Although the positive predictive value was modest (6.4%), this reflects the very low disease prevalence and corresponds to a reasonable number needed to treat of approximately 15. In contrast, the UK-ChiMES-post-LP score did not provide additional diagnostic benefit in our cohort and does not incorporate multiplex PCR data, limiting its relevance in contemporary high-income settings.

Overall, clinical symptoms were largely unspecific across diagnostic categories, and individual laboratory parameters showed limited discriminatory value when assessed in isolation. CRP demonstrated the highest discriminatory accuracy for bacterial meningitis (AUC 0.92), while PCT showed good diagnostic accuracy (AUC 0.85), comparable to pooled estimates from prior meta-analyses (AUC 0.89–0.92) [30, 31]. In contrast, IL-6 showed only moderate diagnostic accuracy in our cohort (AUC 0.78)

This study has several limitations. First, its retrospective single-center design limits generalizability. Second, the very low prevalence of bacterial meningitis reduced statistical power, increasing the influence of individual cases on effect estimates; however, our results are largely consistent with contemporary published literature. Third, the retrospective nature of the analysis is inherently subject to limitations including incomplete or missing data, variability in clinical documentation, potential misclassification or selection bias. Nonetheless, the use of clearly predefined diagnostic categories mitigates some of these limitations. A major strength of this study is the inclusion of a large, unselected real-world cohort of all children undergoing LP for suspected CNS infection, providing a representative reflection of contemporary clinical practice.

In conclusion, in this large, unselected pediatric cohort, bacterial meningitis was rare in the era of routine vaccination and molecular diagnostics, while viral and unresolved infections predominated. Given the limited discriminatory value of clinical features and individual laboratory tests, structured risk stratification is essential. The UK-ChiMES Pre-LP score demonstrated the most favorable diagnostic performance and may safely reduce unnecessary LPs in children evaluated for suspected bacterial meningitis.

## Abbreviations

AUC: Area under the curve
BMS: Bacterial Meningitis Score
CNS: Central nervous system
CRP: C-reactive protein
CSF: Cerebrospinal fluid
EV: Enterovirus
Fig: Figure
HHV6: Human Herpesvirus 6
HSV-1: Herpes simplex virus type 1
ICU: Intensive care unit
IL-6: Interleukin-6
IQR: Interquartile range
LMU: Ludwig Maximilian University
LP: Lumbar puncture
ME: Meningitis/Encephalitis
MRI: Magnetic resonance imaging
NNT: Number needed to test
NPV: Negative predictive value
PCT: Procalcitonin
PCR: Polymerase chain reaction
Pre-LP: Pre-lumbar puncture
Post-LP: Post-lumbar puncture
PPV: Positive predictive value
RBC: red blood cell
ROC: Receiver operating characteristic
TBE: Tick-Borne Encephalitis
VZV: Varicella-zoster virus
WBC: White Blood Cell

## Author contributions

YV, JH, GD, and TS conceived the study. YV, TS, and MMB contributed to the study protocol. YV, BS, JB, MP, AB, and TS extracted and/or clinically evaluated the data. YV and TS performed the formal analysis. YV and TS wrote the first draft of the manuscript. Project administration and supervision were provided by JH, CK, GD, and UB. All authors critically revised the manuscript for important intellectual content, reviewed and approved the final manuscript, had full access to all study data, and had final responsibility for the decision to submit the manuscript for publication.

## Data Availability Statement

The data supporting the findings of this study are not publicly available due to ethical restrictions, as individual-level data sharing is not in accordance with the approved ethics committee vote. Aggregated data may be made available upon reasonable request. Access to anonymized data may be granted after review and approval of a research proposal by the responsible ethics committee. Approved data will be shared through a secure online platform.

## Declaration of generative AI and AI-assisted technologies in the manuscript preparation process

During the preparation of this work the authors used ChatGPT exclusively for language editing and proofreading. After using this tool, the authors reviewed and edited the content as needed and take full responsibility for the content of the published article.

## References

1. Global, regional, and national burden of meningitis and its aetiologies, 1990-2019: a systematic analysis for the Global Burden of Disease Study 2019. Lancet Neurol, 2023. 22(8): p. 685–711.

2. Sadarangani, M., et al., Childhood meningitis in the conjugate vaccine era: a prospective cohort study. Arch Dis Child, 2015. 100(3): p. 292–4.

3. Martin, N.G., et al., Hospital admission rates for meningitis and septicaemia caused by Haemophilus influenzae, Neisseria meningitidis, and Streptococcus pneumoniae in children in England over five decades: a population-based observational study. Lancet Infect Dis, 2014. 14(5): p. 397–405.

4. Rotbart, H.A., Viral meningitis. Semin Neurol, 2000. 20(3): p. 277–92.

5. Martin, N.G., et al., Paediatric meningitis in the conjugate vaccine era and a novel clinical decision model to predict bacterial aetiology. J Infect, 2024. 88(5): p. 106145.

6. van Daalen, K.R., et al., The 2024 Europe report of the <em>Lancet</em> Countdown on health and climate change: unprecedented warming demands unprecedented action. The Lancet Public Health, 2024. 9(7): p. e495–e522.

7. George, B.P., E.B. Schneider, and A. Venkatesan, Encephalitis hospitalization rates and inpatient mortality in the United States, 2000-2010. PLoS One, 2014. 9(9): p. e104169.

8. Iro, M.A., et al., 30-year trends in admission rates for encephalitis in children in England and effect of improved diagnostics and measles-mumps-rubella vaccination: a population-based observational study. Lancet Infect Dis, 2017. 17(4): p. 422–430.

9. Nigrovic, L.E., et al., Clinical prediction rule for identifying children with cerebrospinal fluid pleocytosis at very low risk of bacterial meningitis. JAMA, 2007. 297(1): p. 52–60.

10. Venkatesan, A., et al., Case definitions, diagnostic algorithms, and priorities in encephalitis: consensus statement of the international encephalitis consortium. Clin Infect Dis, 2013. 57(8): p. 1114–28.

11. Dubos, F., et al., Distinguishing between bacterial and aseptic meningitis in children: European comparison of two clinical decision rules. Arch Dis Child, 2010. 95(12): p. 963–7.

12. Nigrovic, L.E., R. Malley, and N. Kuppermann, Meta-analysis of bacterial meningitis score validation studies. Arch Dis Child, 2012. 97(9): p. 799–805.

13. von Elm, E., et al., The Strengthening the Reporting of Observational Studies in Epidemiology (STROBE) statement: guidelines for reporting observational studies. Lancet, 2007. 370(9596): p. 1453–7.

14. Pierart, J. and P. Lepage, [Value of the “Bacterial Meningitis Score” (BMS) for the differential diagnosis of bacterial versus viral meningitis]. Rev Med Liege, 2006. 61(7-8): p. 581–5.

15. Tuerlinckx, D., et al., External validation of the bacterial meningitis score in children hospitalized with meningitis. Acta Clin Belg, 2012. 67(4): p. 282–5.

16. de Ory, F., et al., Viral infections of the central nervous system in Spain: a prospective study. J Med Virol, 2013. 85(3): p. 554–62.

17. Lee, B.E., et al., Paediatric Investigators Collaborative Network on Infections in Canada (PICNIC) study of aseptic meningitis. BMC Infect Dis, 2006. 6: p. 68.

18. Castelblanco, R.L., M. Lee, and R. Hasbun, Epidemiology of bacterial meningitis in the USA from 1997 to 2010: a population-based observational study. Lancet Infect Dis, 2014. 14(9): p. 813–9.

19. Robert, K.-I., Epidemiologisches Bulletin 44/2025: Invasive Meningokokken-Erkrankungen in Deutschland 2024, in Epidemiologisches Bulletin. 2025, Robert Koch-Institut.

20. Agency, U.K.H.S., Invasive meningococcal disease in England: annual laboratory-confirmed cases for epidemiological year 2024 to 2025. 2025, UK Health Security Agency.

21. Kadambari, S., et al., Epidemiological trends in viral meningitis in England: Prospective national surveillance, 2013-2023. J Infect, 2024. 89(3): p. 106223.

22. Kadambari, S., et al., Enterovirus and parechovirus meningitis in infants younger than 90 days old in the UK and Republic of Ireland: a British Paediatric Surveillance Unit study. Arch Dis Child, 2019. 104(6): p. 552–557.

23. de Ory, F., et al., Viral infections of the central nervous system in Spain: a prospective study. J Med Virol, 2013. 85(3): p. 554–62.

24. Kadambari, S., et al., Seven-fold increase in viral meningo-encephalitis reports in England and Wales during 2004-2013. J Infect, 2014. 69(4): p. 326–32.

25. Kohil, A., et al., Viral meningitis: an overview. Arch Virol, 2021. 166(2): p. 335–345.

26. Messacar, K., et al., Encephalitis in US Children. Infect Dis Clin North Am, 2018. 32(1): p. 145–162.

27. Dagostin, F., et al., Ecological and environmental factors affecting the risk of tick-borne encephalitis in Europe, 2017 to 2021. Euro Surveill, 2023. 28(42).

28. Koch-Institut, R., FSME-Risikogebiete in Deutschland (Stand: Januar 2025). Epidemiologisches Bulletin, 2025. 9.

29. Rauer, S., et al., Guidelines for diagnosis and treatment in neurology - Lyme neuroborreliosis. Ger Med Sci, 2025. 23: p. Doc13.

30. Groeneveld, N.S., et al., Biomarkers in paediatric bacterial meningitis: a systematic review and meta-analysis of diagnostic test accuracy. Clin Microbiol Infect, 2025. 31(5): p. 702–712.

31. Kim, H., Y.H. Roh, and S.H. Yoon, Blood Procalcitonin Level as a Diagnostic Marker of Pediatric Bacterial Meningitis: A Systematic Review and Meta-Analysis. Diagnostics (Basel), 2021. 11(5).

