## Supplementary for "Paediatric meningoencephalitis in the molecular diagnostic era: Epidemiological insights from 1,198 suspected cases in Germany between 2016 and 2024"

**Table of Content**

[**Supplementary Table 3.** Bacterial Meningitis Score System [3-5]	8](#_Toc220656381)

[**Supplementary Table 4.** UK-ChiMES-Pre and Post-LP Scores System [1]	9](#_Toc220656382)

### **Supplementary Methods**

##### **Variables of interest and study definitions**

Demographic variables included sex and age at presentation. For analytical purposes, age was categorized into six predefined groups: neonates (0–28 days), infants aged 29 days to <6 months, 6–12 months, 1–4 years, 5–9 years, and 10–17 years. Medical history comprised immunocompromised status or immunosuppressive therapy, underlying malignancy, pre-existing neurological conditions and duration of symptoms prior to hospital presentation. Vaccination status was classified as complete, incomplete, or unknown.

Recorded variables at admission included fever and maximum recorded body temperature within ±48 hours of lumbar puncture (LP), headache (assessed only in children aged ≥4 years), neck stiffness, irritability (in children <4 years of age), rash, vomiting, bulging fontanelle (in infants <9 months of age), altered level of consciousness or personality change lasting more than 24 hours, and seizures occurring within 72 hours before or during hospitalization. In addition, newly onset focal neurological deficits occurring within 72 hours before or during hospitalization were recorded. Antimicrobial therapy was documented for all patients. Clinical outcomes included length of hospital stay, admission to the intensive care unit, initiation of palliative care, and in-hospital mortality. Infections were classified as hospital-acquired if symptom onset occurred more than 48 hours after hospital admission; all remaining cases were considered community-acquired.

Laboratory data included inflammatory and hematologic blood parameters as well as cerebrospinal fluid (CSF) analyses. Blood parameters comprised absolute white blood cell count (WBC) (G/L), absolute neutrophil count (G/L), absolute lymphocyte count (G/L), C-reactive protein (CRP, mg/L), procalcitonin (PCT, ng/mL), and interleukin-6 (IL-6, pg/mL). CSF parameters included corrected WBC count, the proportion of segmented neutrophilic granulocytes (%), lymphocytes (%), and monocytes (%), corrected protein concentration (mg/dL), glucose concentration (mg/dL), and lactate concentration (mmol/L). In cases of traumatic LP predefined correction formulas were applied. CSF leukocyte counts were corrected for erythrocyte contamination using a ratio of 1 × 10⁶/L WBC per 500 × 10⁶/L red blood cells, and measured protein concentrations were corrected by subtracting 1 mg/dL per 1,000 red blood cells/µL.

Imaging and neurophysiological data included neuroimaging and electroencephalographic examinations. Neuroimaging comprised cranial computed tomography, magnetic resonance imaging (MRI), and cranial ultrasound via the anterior fontanelle in infants. All neuroimaging studies were evaluated by a board-certified radiologist with specialization in pediatric radiology, and electroencephalographic recordings were interpreted by a board-certified pediatric neurologist. Each case was reviewed by a pediatric infectious diseases specialist to assess the etiologic relevance of all identified pathogens, including those detected outside the central nervous system (CNS), such as influenza polymerase chain reaction (PCR) from respiratory specimens or Epstein–Barr virus serology. Microbiological findings from non–CSF specimens were interpreted in the context of clinical presentation, laboratory parameters, and CSF results to determine their likelihood of a causal association with the suspected central nervous system infection.

##### **Supplementary Table 1.** STROBE Statement—Checklist

|  | Item No | Recommendation | Page |
| --- | --- | --- | --- |
| **Title and abstract** | 1 | (*a*) Indicate the study’s design with a commonly used term in the title or the abstract | 4 |
|  |  | (*b*) Provide in the abstract an informative and balanced summary of what was done and what was found | 4 |
| Introduction | | |  |
| Background/rationale | 2 | Explain the scientific background and rationale for the investigation being reported | 5 |
| Objectives | 3 | State specific objectives, including any prespecified hypotheses | 5 |
| Methods | | |  |
| Study design | 4 | Present key elements of study design early in the paper | 6 |
| Setting | 5 | Describe the setting, locations, and relevant dates, including periods of recruitment, exposure, follow-up, and data collection | 6 |
| Participants | 6 | (*a*) Give the eligibility criteria, and the sources and methods of selection of participants. Describe methods of follow-up | 6 |
|  |  | (*b*) For matched studies, give matching criteria and number of exposed and unexposed | n/a |
| Variables | 7 | Clearly define all outcomes, exposures, predictors, potential confounders, and effect modifiers. Give diagnostic criteria, if applicable | 6  Supp. 2 |
| Data sources/ measurement | 8* | For each variable of interest, give sources of data and details of methods of assessment (measurement). Describe comparability of assessment methods if there is more than one group | 6, 7,  Supp. 2 |
| Bias | 9 | Describe any efforts to address potential sources of bias | 6, 7 |
| Study size | 10 | Explain how the study size was arrived at | 8 |
| Quantitative variables | 11 | Explain how quantitative variables were handled in the analyses. If applicable, describe which groupings were chosen and why | 6, 7,  Supp. 2 |
| Statistical methods | 12 | (*a*) Describe all statistical methods, including those used to control for confounding | 7 |
|  |  | (*b*) Describe any methods used to examine subgroups and interactions | n/a |
|  |  | (*c*) Explain how missing data were addressed | Table 4. |
|  |  | (*d*) If applicable, explain how loss to follow-up was addressed | n/a |
|  |  | (*e*) Describe any sensitivity analyses | 7 |
| Results | | |  |
| Participants | 13* | (a) Report numbers of individuals at each stage of study—eg numbers potentially eligible, examined for eligibility, confirmed eligible, included in the study, completing follow-up, and analysed | 8 |
|  |  | (b) Give reasons for non-participation at each stage | 8 |
|  |  | (c) Consider use of a flow diagram | Fig. 1 |
| Descriptive data | 14* | (a) Give characteristics of study participants (eg demographic, clinical, social) and information on exposures and potential confounders | Table 2 |
|  |  | (b) Indicate number of participants with missing data for each variable of interest | Table 4 |
|  |  | (c) Summarise follow-up time (eg, average and total amount) | Table 3 |
| Outcome data | 15* | Report numbers of outcome events or summary measures over time | 8, 9  Table 2-4 |
| Main results | 16 | (*a*) Give unadjusted estimates and, if applicable, confounder-adjusted estimates and their precision (eg, 95% confidence interval). Make clear which confounders were adjusted for and why they were included | Table 2-4 |
|  |  | (*b*) Report category boundaries when continuous variables were categorized | Table 2 |
|  |  | (*c*) If relevant, consider translating estimates of relative risk into absolute risk for a meaningful time period | n/a |
| Other analyses | 17 | Report other analyses done—eg analyses of subgroups and interactions, and sensitivity analyses | 9 |
| Discussion | | |  |
| Key results | 18 | Summarise key results with reference to study objectives | 10, 11 |
| Limitations | 19 | Discuss limitations of the study, taking into account sources of potential bias or imprecision. Discuss both direction and magnitude of any potential bias | 11 |
| Interpretation | 20 | Give a cautious overall interpretation of results considering objectives, limitations, multiplicity of analyses, results from similar studies, and other relevant evidence | 11 |
| Generalisability | 21 | Discuss the generalisability (external validity) of the study results | 11, 12 |
| Other information | | |  |
| Funding | 22 | Give the source of funding and the role of the funders for the present study and, if applicable, for the original study on which the present article is based | 13 |

##### **Supplementary Figure 1.** Definition of clinical phenotypes

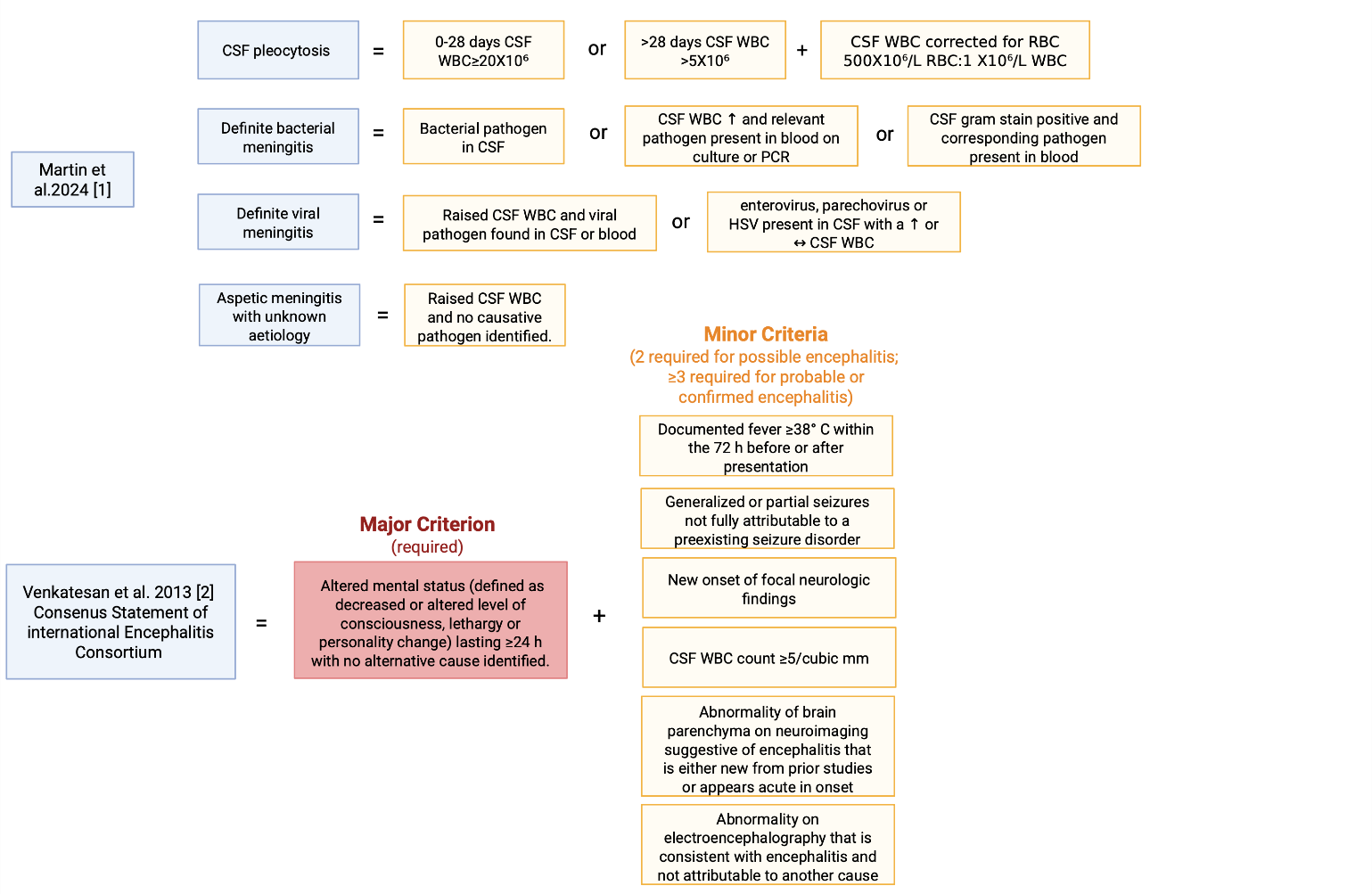

##### **Supplementary Table 2.** Definition of clinical phenotypes

| Definitions |  | Sources |
| --- | --- | --- |
| CSF pleocytosis | A raised CSF WBC count (CSF pleocytosis) was defined as age 0-28 days CSF WBC ≥20 X10^6^/L, age >28 days CSF WBC >5 X10^6^/L. The CSF white blood cell (WBC) count was corrected for red blood cell count at a ratio of 500X10^6^/L RBC:1 X10^6^/L WBC. | [1] |
| Definite bacterial meningitis | Bacterial pathogen in CSF, or raised CSF WBC and relevant pathogen present in blood on culture or PCR, or CSF gram stain positive and corresponding pathogen present in blood. Relevant pathogens in blood included *N. meningitidis, S. pneumoniae, H. influenzae,* Group B Streptococcus, *E. coli*, *S. aureus* and Group A Streptococcus. | [1] |
| Definite viral meningitis | Raised CSF WBC and viral pathogen found in CSF or blood; or enterovirus, parechovirus or HSV present in CSF with a raised or normal CSF WBC. | [1] |
| Aseptic meningitis with unknown aetiology | Raised CSF WBC and no causative pathogen identified. | [1] |
| Possible encephalitis,  probable encephalitis | Major Criterion (required): Patients presenting to medical attention with altered mental status (defined as decreased or altered level of consciousness, lethargy or personality change) lasting ≥24 h with no alternative cause identified  Minor Criteria (2 required for possible encephalitis, ≥3 required for probable encephalitis):  Documented fever ≥38° C (100.4°F) within the 72 h before or after presentation, generalized or partial seizures not fully attributable to a preexisting seizure disorder, new onset of focal neurologic findings, CSF WBC count ≥5/cubic mm, Abnormality of brain parenchyma on neuroimaging suggestive of encephalitis that is either new from prior studies or appears acute in onset, Abnormality on electroencephalography that is consistent with encephalitis and not attributable to another cause | [2] |
| Confirmed encephalitis | Confirmed encephalitis was defined as fulfillment of all criteria for probable encephalitis, with additional microbiological confirmation of a causative pathogen | [2] |
| Diagnosis of a complex febrile seizure | All patients who were discharged home with a diagnosis of a complex febrile seizure in their discharge letter. |  |
| CNS Lyme  disease | Patients with neurological symptoms who underwent lumbar puncture and had serologically confirmed Borrelia infection |  |
| Control Group | All patients without pleocytosis, without pathogen detection in CSF multiplex PCR, and without a diagnosis of possible or probable encephalitis |  |

##### **Supplementary Table 3.** Bacterial Meningitis Score System [3-5]

**Components of the Bacterial Meningitis Score**

Positive CSF Gram stain

CSF absolute neutrophil count ≥1000 cells/µL

CSF protein ≥80 mg/dL

Peripheral blood absolute neutrophil count ≥10 000 cells/µL

History of seizure before or at the time of presentation

Patients are classified as very low risk if none of these variables are present. Patients were classified as not very low risk if at least one of these variables was present.

##### **Supplementary Table 4.** UK-ChiMES-Pre and Post-LP Scores System [1]

| Variables | Score |
| --- | --- |
| **Pre-LP** | |
| Vomiting | 0.5 |
| Altered consciousness | 0.5 |
| Bulging fontanelle | 2 |
| Lymphocyte count <2.4 x10^9^/L | 1.5 |
| CRP >50.9 mg/L^*^ | 2.5 |
| **Post-LP** | |
| Rash |  |
| Non-blanching | 1.5 |
| Other | 0 |
| No rash | 0.5 |
| Lymphocyte count <2.9 x10^9^/L | 1 |
| CRP >50.5 mg/L | 2.5 |
| CSF glucose <2.15 mmol/L | 1.5 |
| CSF protein >1.15 g/L | 1.0 |
| CSF WBC > 187.5 x10^6^/L | 1.0 |
| CSF neutrophils > 66.25 x10^6^/L | 1.0 |

A positive Pre-LP-score required >2 points; a positive post LP-Score required >4 points in the respective scoring system

### **Supplementary Results**

##### **Supplementary Figure 2.** ROC curves of CRP, PCT and IL-6 for Definite Bacterial Meningitis

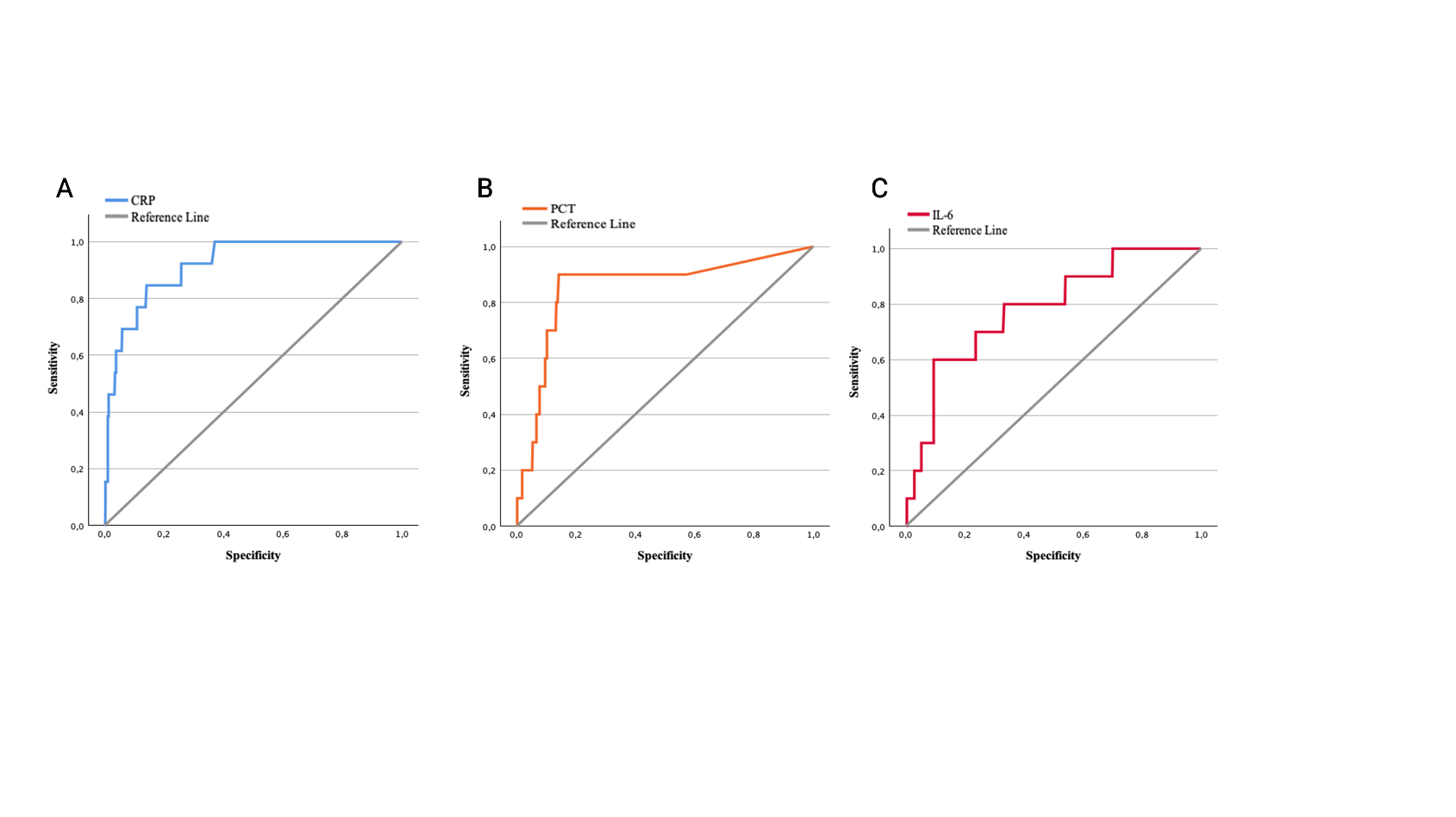

Reiceiver Operator Characteristics (ROC) curves to detect bacterial meningitis are shown for C-reactive Protein (A), Procalcitonin (B) and Interleukin-6 (C)

##### **Supplementary Table 5.** ROC characteristics of CRP, PCT and IL-6 for Definite Bacterial Meningitis

| Test Result Variable | Area | Std. Error | Asymptotic Sig. | 95% CI Lower Bound | 95% CI Upper Bound | Cutt-off (Youden) | Sensitivity | Specificity | Youden index |
| --- | --- | --- | --- | --- | --- | --- | --- | --- | --- |
| CRP | 0.919 | 0.030 | 0.000 | 0.859 | 0.979 | ≥5.07 mg/dL | 84.6% | 85.9% | 0.705 |
| PCT | 0.852 | 0.070 | 0.000 | 0.714 | 0.990 | ≥3.35 ng/mL | 90.0% | 85.8% | 0.758 |
| IL-6 | 0.782 | 0.072 | 0.000 | 0.641 | 0.923 | ≥949 pg/mL | 60.0% | 90.5% | 0.50 |

##### **Supplementary Table 6.** Missed cases of confirmed bacterial meningitis: Scoring table

|  | Pre-LP >2 | Post-LP >4 | BMS ≥1 (≥29 Days) |
| --- | --- | --- | --- |
| P1 | 4.5 | 8 | 3 |
| P2 | 5 | 6.5 | 1 |
| P3 | 4 | 5 |  |
| P4 | 4.5 | 8.5 | 1 |
| P5 | 5 | 8.5 |  |
| P6 | 2.5 | 3 | 1 |
| P7 | 2.5 | 7.5 |  |
| P8 | 2.5 | 5 | 0 |
| P9 | 3.5 | 3 | 3 |
| P10 | 4.5 | 7 | 1 |
| P11 | 4.5 | 8.5 | 4 |
| P12 | 0 | 2.5 | 0 |
| P13 | 2 | 6 | 2 |

Patient P1-P13 are shown with their corresponding scores. A positive UK-ChiMES Pre-LP-score required >2 points and a positive Post-LP-score required >4 points. Patients with a BMS ≥1 point are considered not very low risk (Supplementary Tables 3 and 4). Missed cases of confirmed bacterial meningitis are shown in grey.

##### **Supplementary Table 7.** Missed cases of confirmed bacterial meningitis: Case descriptions

|  | Failed risk score | Age | Hospital stay | Diagnosis | Description | Comment |
| --- | --- | --- | --- | --- | --- | --- |
| P6 | Post-LP | 6 weeks | 14 days | Neonatal sepsis caused by Streptococcus pneumoniae | Markedly hemorrhagic CSF; corrected WBC count of 70; neurologically unremarkable | Potentially false positive pleocytosis after predefined correction of 500 × 10⁶/L RBC to 1 × 10⁶/L WBC.  Not considered to be a meningitis by treating physicians in original discharge note. |
| P8 | BMS | 11 months | 24 days | Urosepsis caused by Escherichia coli | CSF WBC count 6 cells/µl; blood and urine cultures positive for Escherichia coli; former extremely preterm infant (GA 26 + 2 weeks) | CSF WBC just above predefined threshold, but unusually low for bacterial meningitis. Not considered to be a meningitis by treating physicians in original discharge note. |
| P9 | Post-LP | 10 months | 28 days | Fulminant sepsis due to Streptococcus pneumoniae with pyogenic meningitis | Altered mental status with seizures and focal neurological deficits; intubated due to status epilepticus; WBC count 37 cells/µl |  |
| P12 | Pre-/post-LP, BMS | 6 months | 11 days | HHV6 meningitis and pneumococcal bacteremia | Markedly hemorrhagic CSF (with a corrected WBC count of 76 cells/µl; good overall clinical condition; neurologically unremarkable | Potentially false positive pleocytosis after predefined correction of 500 × 10⁶/L RBC to 1 × 10⁶/L WBC. Not considered to be a meningitis by treating physicians in original discharge note. |
| P13 | Pre-LP | 7 years | 12 days | Otitis media followed by mastoiditis, caused by Streptococcus pyogenes | Significantly hemorrhagic CSF with a corrected WBC count of 1,843; stable overall condition; initially neck pain and vomiting |  |

1.	Martin, N.G., et al., *Paediatric meningitis in the conjugate vaccine era and a novel clinical decision model to predict bacterial aetiology.* J Infect, 2024. **88**(5): p. 106145.

2.	Venkatesan, A., et al., *Case definitions, diagnostic algorithms, and priorities in encephalitis: consensus statement of the international encephalitis consortium.* Clin Infect Dis, 2013. **57**(8): p. 1114-28.

3.	Dubos, F., et al., *Distinguishing between bacterial and aseptic meningitis in children: European comparison of two clinical decision rules.* Arch Dis Child, 2010. **95**(12): p. 963-7.

4.	Nigrovic, L.E., R. Malley, and N. Kuppermann, *Meta-analysis of bacterial meningitis score validation studies.* Arch Dis Child, 2012. **97**(9): p. 799-805.

5.	Nigrovic, L.E., et al., *Clinical prediction rule for identifying children with cerebrospinal fluid pleocytosis at very low risk of bacterial meningitis.* JAMA, 2007. **297**(1): p. 52-60.

### **References**
